# Measuring College Student Attitudes Toward COVID-19 Vaccinations

**DOI:** 10.1101/2021.10.30.21265699

**Authors:** Z.W. Taylor, Ibrahim Bicak, Joshua Childs, Carla Fletcher, Allyson Cornett

## Abstract

This survey explores attitudes of 1,197 currently enrolled college students regarding their comfort taking a COVID-19 vaccine. Results suggest most college students are willing to take a COVID-19 vaccine if their institution requires it to return to campus in subsequent semesters. However, certain students of Color, students with disabilities, and adult students may be less willing to take a COVID-19 vaccine if it were required before or during an on-campus semester. Finally, many college students do not understand that COVID-19 vaccines will be free, possibly affecting these students’ willingness to vaccinate and their perceptions of safely and affordably returning to campus. Implications for postsecondary policy and leadership are addressed.

---

After March 11th, 2020, when the World Health Organization (WHO) declared COVID-19 (coronavirus) a global pandemic (World Health Organization, 2020), colleges and universities briskly moved students, faculty, and staff online in an abundance of safety and caution. This move to online learning was particularly burdensome, both in terms of human and financial capital, for institutions with large on campus populations. Moving students online also removed them from residence halls and student affairs related activities on campus, many of which employ thousands of higher education professionals across the United States and the world (Hubler, 2020). Additionally, on campus activities, such as becoming involved in student organizations and socializing with classmates in true in person settings have been proven to increase retention and graduation rates, compounding the difficulty of a shift to online learning (Bawa, 2016).

Given the many hurdles presented by COVID-19, many institutions of higher education are exploring how to reopen their doors in subsequent semesters. Reopening would not only bring students back to campus but also bring faculty and staff back to campus and re-employ thousands of laid off or furloughed workers across the United States and the World (Hubler, 2020). As has been well-documented across dozens of news outlets, the development of a viable, safe, and effective COVID-19 vaccine became a reality in early 2021, as both Pfizer and Moderna mRNA vaccines cleared emergency authorization and were available to certain populations (Centers for Disease Control and Prevention, 2021). The public availability of vaccines in early 2021 was often limited to high-risk populations including those who are 65 and older or those who were essential workers working in healthcare, such as hospital and clinic workers (Centers for Disease Control and Prevention, 2021). However, as the COVID-19 vaccine becomes more widely available, colleges and universities, especially those with robust on campus living facilities and residence halls, maybe face difficult questions:

1. How comfortable do college students feel taking a COVID-19 vaccine?
2. Would college students take a COVID-19 if it were required by their institution to return to campus?
3. Mid-semester, would postsecondary students withdraw from classes if their institution required a COVID-19 vaccine to stay on campus?
4. Do postsecondary students understand that COVID-19 vaccines will be free, either through insurance or federal emergency funds, per U.S. Government mandates (U.S. Department of Health and Human Services, 2020)?

These are the questions that this brief answers through a survey of 1,197 currently-enrolled postsecondary students in the United States at all three levels: two-year students, four-year students, and graduate students. Results suggest many postsecondary students are willing to take a COVID-19 vaccine if their institution required it to return to on-campus learning, however some students may be wary of returning to campus without a vaccine only to be told mid-semester that a vaccine is required to stay on-campus. Moreover, many postsecondary students may not know that COVID-19 vaccines will be free, potentially alerting institutions to the need for clear communication with students regarding COVID-19 vaccine procedures and cost.

## Methods

Data for this survey were gathered in January 2021 when public availability of COVID-19 vaccines became clearer through official communication from the Centers for Disease Control and the World Health Organization (Centers for Disease Control and Prevention, 2021). The research team employed Amazon Mechanical Turk (MTurk) to survey postsecondary students currently enrolled at institutions of higher education. MTurk has been found to be a unique and robust source of human intelligence services, including survey completion in educational contexts (Follmer et al., 2017). Several recent studies in education focused on financial aid jargon (Taylor & Bicak, 2019) and computer science education (Hellas et al., 2020) have used MTurk to answer research questions that require a large, nationally representative dataset, akin to the study at hand related to postsecondary student attitudes toward COVID-19 vaccinations.

The survey asked for a student’s birth year, race, gender, first generation in college status (defined as neither parent earning any level of postsecondary credential), self-reported dis/ability status, educational level (two-year, four-year, or graduate), enrollment status (part- or full-time), and current mode of education (on-campus, online, or hybrid). A geospatial map of the location of survey respondents (n=1,197) can be found in Figure 1 below:

**Figure 1.**
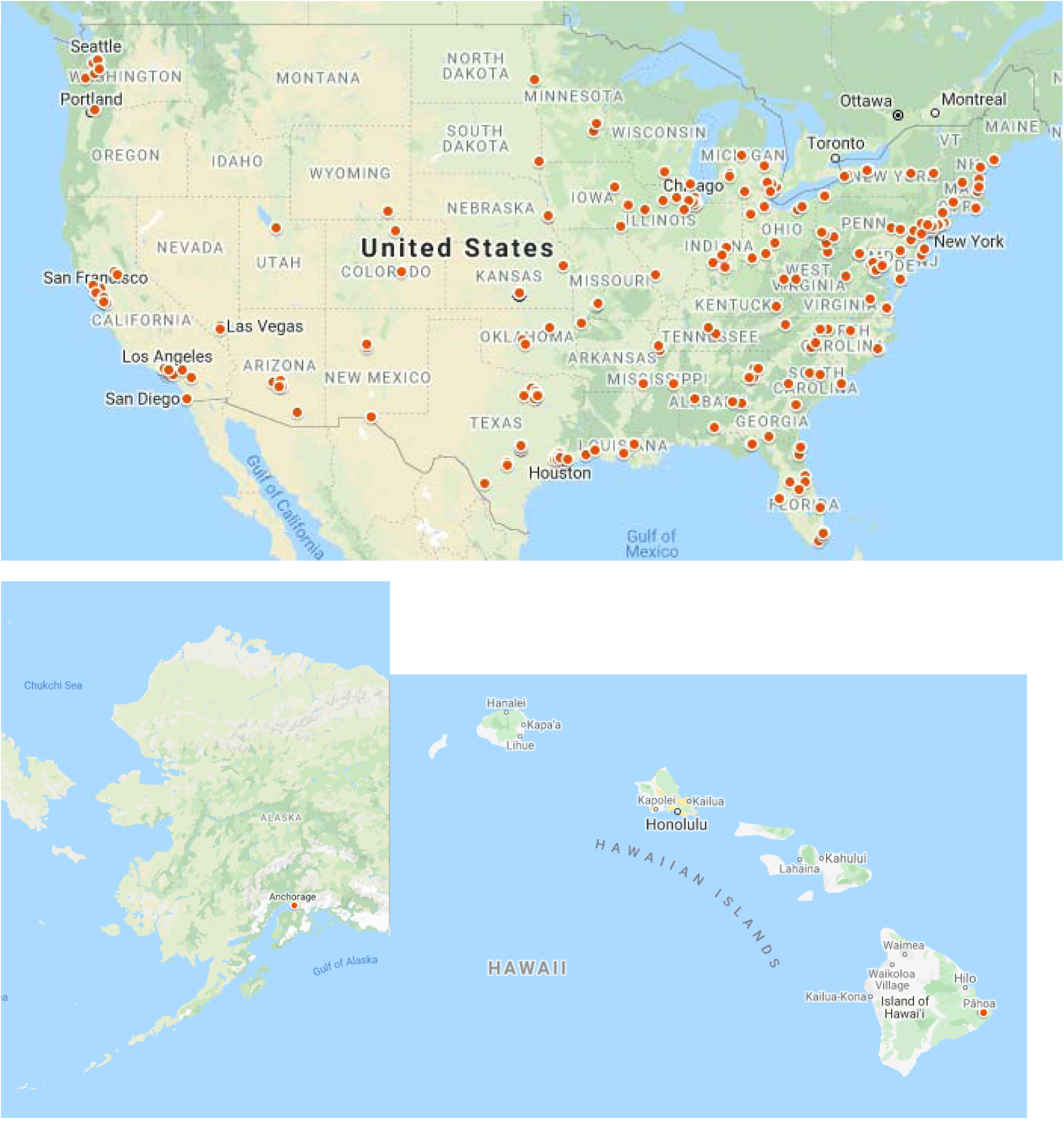
Geospatial map of survey participants (n=1,197)

Then, students were asked to report their level of comfort regarding taking a COVID-19 vaccine on a seven-point Likert scale, followed by three follow-up questions:

1. Would you take a COVID-19 vaccine if your school REQUIRED it for you to return to campus? (Yes/No)
2. Mid-semester, would you withdraw from classes at your school if your school REQUIRED a COVID-19 vaccine to stay on campus? (Yes/No)
3. How much will the COVID-19 vaccine cost you? (Nothing - the vaccine will be free through insurance or the federal government/$1-99/$100-199/$200+)

### Limitations

As with any survey study, this study is limited primarily by the reliability and validity of the survey data. This study gathered data from MTurk and participants who self-reported their college enrollment status as well as other demographics. Moreover, this study is also limited by its temporal nature, meaning that college student attitudes toward COVID-19 vaccines may drastically change over time as vaccine efficacy is reported and vaccines become safer and more available to the general public. In addition, as institutions of higher education release their reopening plans for full on-campus immersion in the 2021 and 2022 academic years, college student attitudes towards taking a COVID-19 vaccine may also change due to idiosyncratic institutional planning. Yet, the strengths of this study is its sample size (n=1,197), rendering it robust for quantitative analysis and generalizability, while also reporting timely and critical data for institutions of higher education: For these reasons, the research team feels the study’s strengths outweigh its limitations.

## Results

Descriptive statistics of survey responses by demographic can be found in Table 1 below:

**Table 1.**
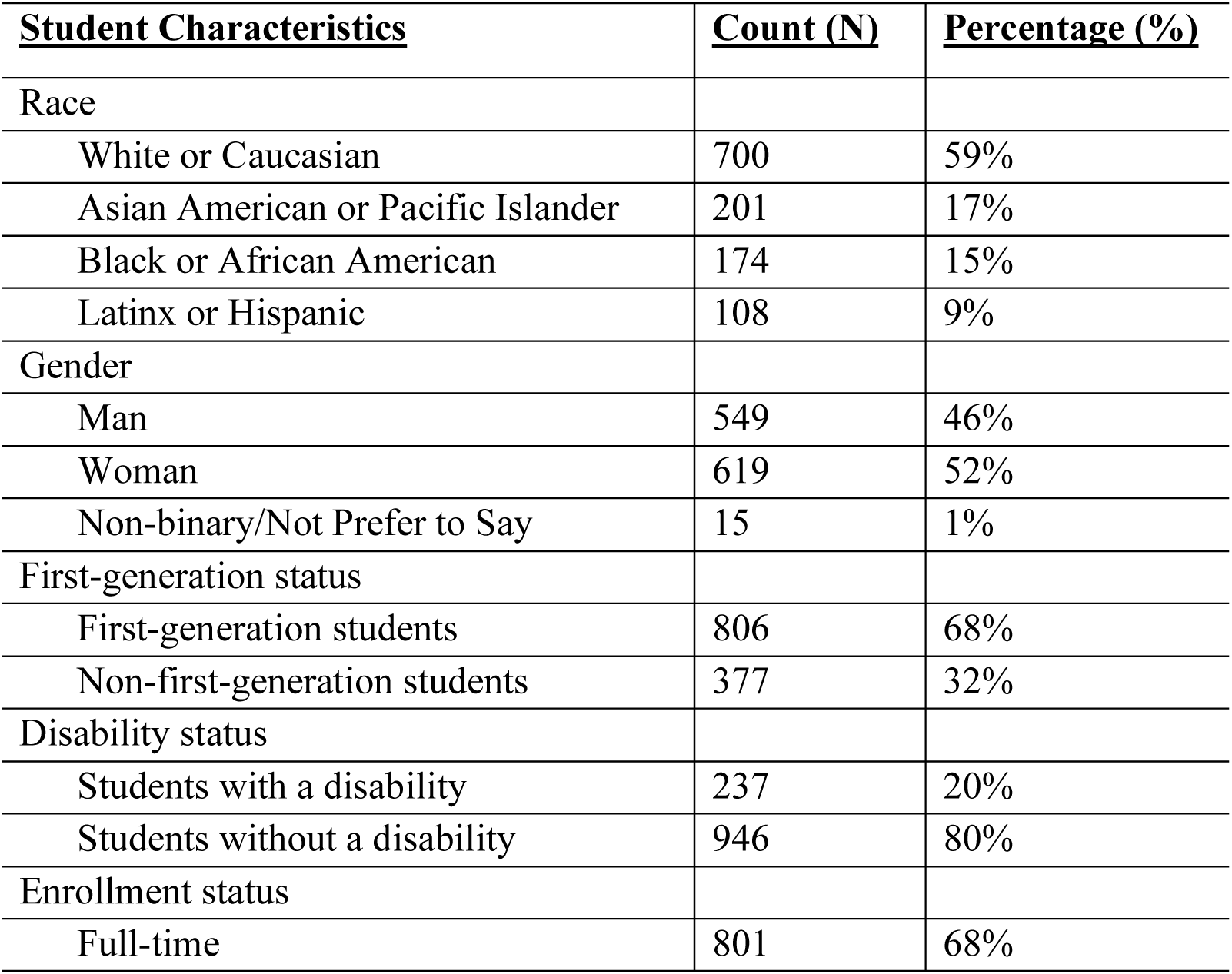
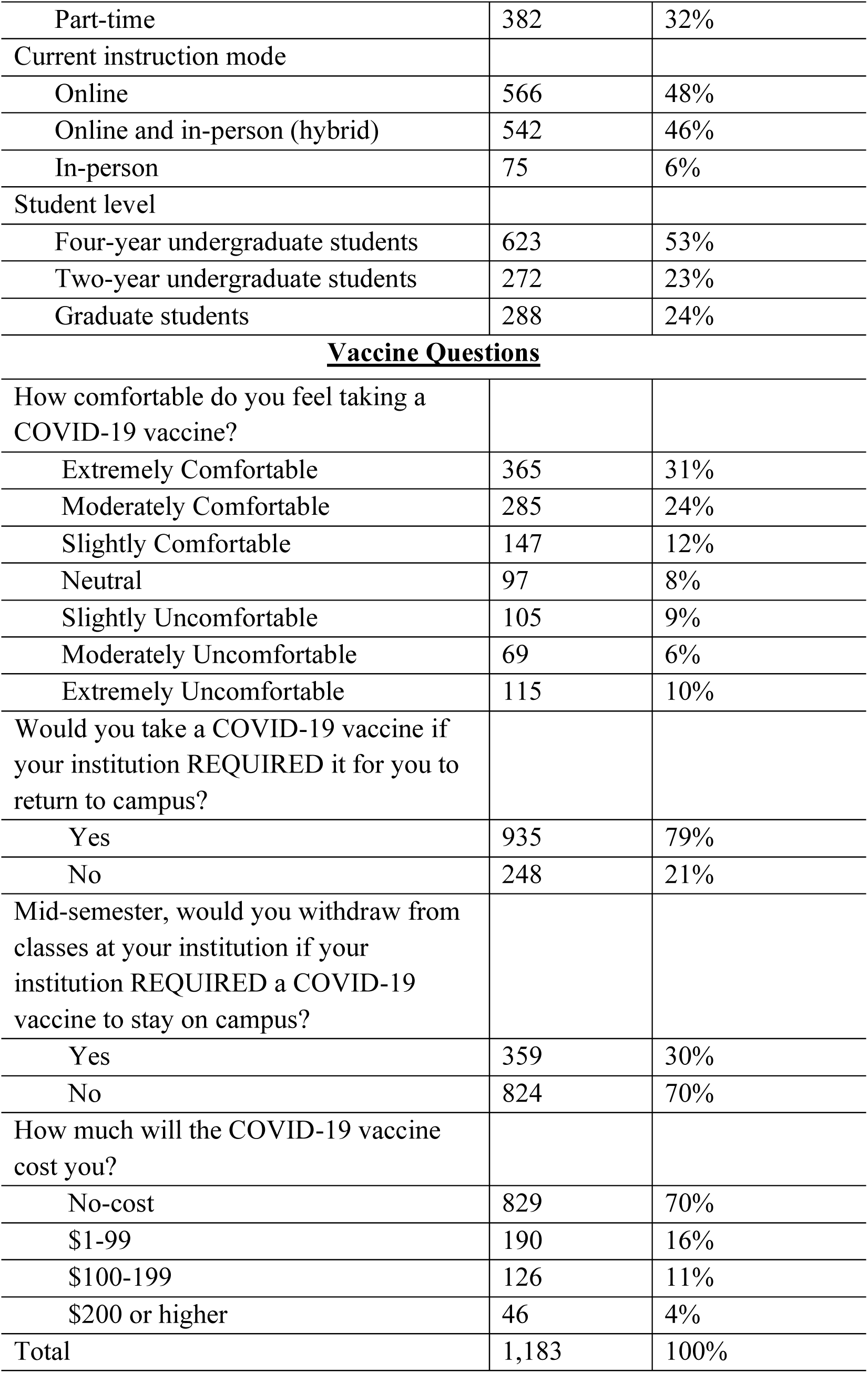
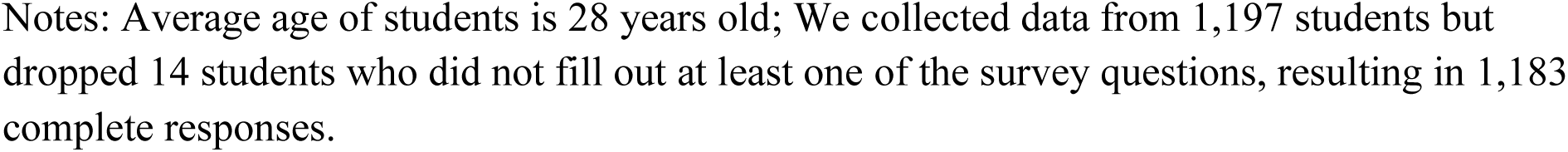
Descriptive statistics of survey sample and responses (n=1,183)

| **<u>Student Characteristics</u>** | **<u>Count (N)</u>** | **<u>Percentage (%)</u>** |
| --- | --- | --- |
| Race |  |  |
| White or Caucasian | 700 | 59% |
| Asian American or Pacific Islander | 201 | 17% |
| Black or African American | 174 | 15% |
| Latinx or Hispanic | 108 | 9% |
| Gender |  |  |
| Man | 549 | 46% |
| Woman | 619 | 52% |
| Non-binary/Not Prefer to Say | 15 | 1% |
| First-generation status |  |  |
| First-generation students | 806 | 68% |
| Non-first-generation students | 377 | 32% |
| Disability status |  |  |
| Students with a disability | 237 | 20% |
| Students without a disability | 946 | 80% |
| Enrollment status |  |  |
| Full-time | 801 | 68% |
| Part-time | 382 | 32% |
| Current instruction mode |  |  |
| Online | 566 | 48% |
| Online and in-person (hybrid) | 542 | 46% |
| In-person | 75 | 6% |
| Student level |  |  |
| Four-year undergraduate students | 623 | 53% |
| Two-year undergraduate students | 272 | 23% |
| Graduate students | 288 | 24% |
| **Vaccine Questions** |  |  |
| How comfortable do you feel taking a COVID-19 vaccine? |  |  |
| Extremely Comfortable | 365 | 31% |
| Moderately Comfortable | 285 | 24% |
| Slightly Comfortable | 147 | 12% |
| Neutral | 97 | 8% |
| Slightly Uncomfortable | 105 | 9% |
| Moderately Uncomfortable | 69 | 6% |
| Extremely Uncomfortable | 115 | 10% |
| Would you take a COVID-19 vaccine if your institution REQUIRED it for you to return to campus? |  |  |
| Yes | 935 | 79% |
| No | 248 | 21% |
| Mid-semester, would you withdraw from classes at your institution if your institution REQUIRED a COVID-19 vaccine to stay on campus? |  |  |
| Yes | 359 | 30% |
| No | 824 | 70% |
| How much will the COVID-19 vaccine cost you? |  |  |
| No-cost | 829 | 70% |
| \$1-99 | 190 | 16% |
| \$100-199 | 126 | 11% |
| \$200 or higher | 46 | 4% |
| Total | 1,183 | 100% |

Notes: Average age of students is 28 years old; We collected data from 1,197 students but dropped 14 students who did not fill out at least one of the survey questions, resulting in 1,183 complete responses.

### Descriptive statistics of survey sample and responses (n=1,183)

Notably, 68 percent of these college students identified as first-generation students, and one in five self-disclosed some type of disability. While 46 percent of respondents identified as men and 52 percent as women, one percent indicated they were non-binary or preferred not to say. When surveyed students were asked to identify the current mode of instruction received, 48 percent reported they were only receiving online instruction, 46 percent selected a hybrid model of instruction with a mix of in-person and online learning, and six percent answered they were only receiving in-person instruction. Part-time students were also well-represented in the sample; nearly a third of respondents (32 percent) were enrolled at their institution part-time. The average age of students in the survey was 28 years old. See Table 1 for more demographic characteristics of students in the sample.

Generally, students expressed comfort with the idea of taking a COVID-19 vaccine, with 67 percent of students reporting feeling either slightly comfortable (12 percent), moderately comfortable (24 percent), or extremely comfortable (31 percent); however, a quarter of surveyed students were uncomfortable to some degree, with 10 percent reporting they were extremely uncomfortable with the idea. Seventy-nine percent of these students supported vaccinations before returning to campus, with only 21 percent indicating otherwise. Further, 30 percent of respondents would withdraw from classes at their institution if a COVID-19 vaccine was required to stay on campus. When queried on how much the COVID-19 vaccine would cost them, 70 percent of students expected there would be no-cost, 16 percent believed it would cost $1-$99, 11 percent thought the cost would be $100-$199, and four percent said the vaccine would cost them over $200.

Regression analyses predicting college students’ levels of comfort with COVID-19 vaccines (Model 1), attitudes towards returning to campus (Models 2 and 3), and vaccination costs (Model 4) can be found in Table 2 below:

**Table 2.** Regression analyses predicting college students’ COVID-19 vaccine attitudes (n=1,183)

|  | (1) | (2) | (3) |
| --- | --- | --- | --- |
| Variable | Vaccine Comfort | Take Vaccine | Withdraw |
| Race (Control=White) |  |  |  |
| Asian American or Pacific Islander | 0.067<br>(0.038) | 0.033<br>(0.034) | -0.062<br>(0.034) |
| Black or African American | -0.157***<br>(0.038) | -0.005<br>(0.034) | -0.097*<br>(0.034) |
| Latinx or Hispanic | -0.068<br>(0.047) | 0.061<br>(0.042) | -0.053<br>(0.042) |
| Gender (Control=Man) |  |  |  |
| Woman | -0.150***<br>(0.028) | -0.078*<br>(0.024) | 0.056*<br>(0.025) |
| Non-binary/not disclosed | 0.220<br>(0.119) | 0.163<br>(0.105) | -0.281*<br>(0.106) |
| Adult (Control=Young) |  |  |  |
| 25 or older | 0.033<br>(0.029) | -0.054*<br>(0.026) | 0.091***<br>(0.026) |
| First-gen | 0.197***<br>(0.029) | 0.139***<br>(0.026) | 0.029<br>(0.026) |
| Student with a disability | -0.055<br>(0.035) | -0.048<br>(0.031) | 0.323***<br>(0.031) |
| Full-time students | 0.022<br>(0.030) | 0.070*<br>(0.027) | -0.000<br>(0.027) |
| Instruction Mode (Control=Online) |  |  |  |
| Hybrid | -0.011<br>(0.028) | -0.025<br>(0.025) | 0.031<br>(0.025) |
| In-person | 0.081<br>(0.056) | -0.058<br>(0.049) | -0.057<br>(0.050) |
| Student level (Control=four-year) |  |  |  |
| Two-year students | -0.007<br>(0.034) | -0.061*<br>(0.030) | 0.023<br>(0.030) |
| Graduate students | -0.020 | 0.059* | -0.018 |
|  | (0.034) | (0.030) | (0.030) |
| Vaccine cost (Reference= No cost) | 0.028 | -0.002 | 0.295*** |
|  | (0.031) | (0.027) | (0.027) |
| Constant | 0.608*** | 0.732*** | 0.072 |
|  | (0.050) | (0.044) | (0.045) |
| Observations | 1,183 | 1,183 | 1,183 |
| R-squared | 0.101 | 0.075 | 0.261 |
| Notes: Standard errors in parentheses; *** p<0.001, ** p<0.001, * p<0.05 |  |  |  |

Results from table 2 show that female students reported feeling less comfortable with receiving the vaccine (p<.001) and were less likely to say they would take the vaccine if it was required by their institution (p<.05) as compared to male students. Additionally, female (p<.05) and non-binary students, as well as students who did not disclose their gender (p<.05), were more likely to say they would withdraw from classes if the vaccine was required by their institution (p<.05). Similarly, students aged 25 and older were less likely to commit to taking the vaccine if it were a requirement by the institution (p<.05) and more likely to report they would withdraw from classes if it were required (p<.001) as compared to younger students. Black/African American students reported feeling less comfortable with receiving the vaccine (p<.001) and were more likely to say they would withdraw from the institution mid-semester if a vaccination was required to remain enrolled (p<.05), compared to White students. White students and Black/African American students were not significantly different in their commitment to taking the vaccine if it were a requirement by the institution. There were no significant differences in survey responses between White students and Asian American/Pacific Islander students or Latinx/Hispanic students.

First generation students (p<.001) were more likely to express comfort with receiving the vaccine (p<.001) and more likely to say they would take the vaccine if it was required by their institution (p<.001), but there was no statistically significant difference between first generation and non-first generation students regarding withdrawing from classes if vaccination were required. Students attending full time were also more likely to say they would take the vaccine if it was required (p<.05), compared to students not attending full-time. Using students attending four-year institutions as the control, graduate students (p<.05) were more likely to report that they would take the vaccine if it were required by their school, while students attending two-year institutions were less likely to say so (p<.05).

Students with a disability (p<.001) and students who believed they would need to pay to be vaccinated (p<.001) were more likely to say they would withdraw from classes mid-semester if their institution required vaccination, but neither group showed any significant differences from their comparison group in reporting their comfort with the vaccine or their willingness to take the vaccine if required. There were no significant differences in responses about comfort level with the vaccine, willingness to take the vaccine, or likelihood of withdrawing based on whether the student attended classes fully online, fully in-person, or a hybrid of online and in-person.

## Discussion and Implications for Policy and Leadership

Central to the conversation about universities’ Fall 2021 and beyond opening plans has focused on vaccine distribution. Many communications from higher education officials has focused on the necessity for vaccines to be widely available and accessible to the staff, students, and faculty that make up university communities. While COVID-19 variants and community surges are to be expected in the foreseeable future, the progress of vaccines and the potential to inoculate entire student bodies and personnel presents promise. However, vaccine shortages, modes of delivery, and individual willingness to become vaccinated remain an ongoing hurdle for higher education officials. The distribution of the COVID-19 vaccine has fallen on state and local officials, who are often competing with one another to procure a limited supply. This has not only created a tumultuous vaccine rollout, but misalignment between federal, state, and local efforts to vaccinate the public. This has put higher education institutions in precarious positions, where many serve as vaccine distribution hubs, yet must consider their role in the spread of COVID-19 if they were to fully return to in-person operations. However, there are promising policy approaches that universities could embark on in order to help advance COVID-19 vaccine distribution and ensuring that their campuses remain safe for students, staff, and faculty.

Because COVID-19 transmission is more likely to occur in densely populated communities near university campuses, vaccinating students has become a top priority for university officials. However, and as we sought to discover in this study, requiring students to become vaccinated before returning to campus is an important empirical question that needs to be fully addressed through efficient policies and practices that promotes the safety of the university community. Our findings showed that students supported vaccinations before returning to campus, and universities are well positioned to provide guidance on the importance of vaccines on affecting community safety and a return to normal operations. Universities could leverage the typical expectations of students’ college experiences with the necessity of getting vaccinated, and the possible end of social distancing policies, mask mandates, and stay-at-home orders. Also, universities should centralize messaging around COVID-19 vaccines to reduce misinformation on their effectiveness, confusion over insurance policies and coverage, and confusion on vaccine eligibility and availability.

Ultimately, it is critical for postsecondary institutions to communicate COVID-19 vaccination policy is clearly and concisely to all students, especially minoritized students of color, students with disabilities, and vulnerable adult populations to ensure that vaccine procedures and costs are not a deterrent from being vaccinated. If colleges and universities across the United States and the World want to safely open campuses in Fall 2021 and beyond, clear communication of often complex medical information is necessary to ensure students, faculty, and staff that COVID-19 vaccines are safe, affordable, and indeed necessary for the health of the public and the health of postsecondary campuses now and into the future.

## Data Availability

All data produced in the present study are available upon reasonable request to the authors.

## Notes

### Competing Interest Statement

The authors have declared no competing interest.

### Funding Statement

This study did not receive any funding.

### Author Declarations

Institutional review board of Texas State University gave ethical approval for this work in 2020.

